# Predicting Autism Spectrum Disorder: Transformer-Based Deep Learning Ensemble Framework Using Health Administrative & Birth Registry Data

**DOI:** 10.1101/2024.07.03.24309684

**Authors:** Kevin Dick, Emily Kaczmarek, Robin Ducharme, Alexa C. Bowie, Alysha L.J. Dingwall-Harvey, Heather Howley, Steven Hawken, Mark C. Walker, Christine M. Armour

## Abstract

**Background:** Early diagnosis and access to resources, support and therapy are critical for improving long-term outcomes for children with autism spectrum disorder (ASD). ASD is typically detected using a case-finding approach based on symptoms and family history, resulting in many delayed or missed diagnoses. While population-based screening would be ideal for early identification, available screening tools have limited accuracy. This study aims to determine whether machine learning models applied to health administrative and birth registry data can identify young children (aged 18 months to 5 years) who are at increased likelihood of developing ASD.

**Methods:** We assembled the study cohort using individually linked maternal-newborn data from the Better Outcomes Registry and Network (BORN) Ontario database. The cohort included all live births in Ontario, Canada between April 1st, 2006, and March 31st, 2018, linked to datasets from Newborn Screening Ontario (NSO), Prenatal Screening Ontario (PSO), and Canadian Institute for Health Information (CIHI) (Discharge Abstract Database (DAD) and National Ambulatory Care Reporting System (NACRS)). The NSO and PSO datasets provided screening biomarker values and outcomes, while DAD and NACRS contained diagnosis codes and intervention codes for mothers and offspring. Extreme Gradient Boosting models and large-scale ensembled Transformer deep learning models were developed to predict ASD diagnosis between 18 and 60 months of age. Leveraging explainable artificial intelligence methods, we determined the impactful factors that contribute to increased likelihood of ASD at both an individual- and population-level.

**Results:** The final study cohort included 703,894 mother-offspring pairs, with 10,964 identified cases of ASD. The best-performing ensemble of Transformer models achieved an area under the receiver operating characteristic curve of 69.6% for predicting ASD diagnosis, a sensitivity of 70.9%, a specificity of 56.9%. We determine that our model can be used to identify an enriched pool of children with the greatest likelihood of developing ASD, demonstrating the feasibility of this approach.

**Conclusions:** This study highlights the feasibility of employing machine learning models and routinely collected health data to systematically identify young children at high likelihood of developing ASD. Ensemble transformer models applied to health administrative and birth registry data offer a promising avenue for universal ASD screening. Such early detection enables targeted and formal assessment for timely diagnosis and early access to resources, support, or therapy.

## 1. Introduc0on

Autism spectrum disorder (ASD) is a neurodevelopmental disorder characterized by enduring difficulties in social interaction, speech and nonverbal communication, and repetitive behaviors ^1^. Individuals with ASD may be at increased risk for experiencing stressful and traumatic life events, the sequelae of which can negatively impact mental health through the development of comorbid psychopathology and/or worsening of the core symptoms of ASD ^2,3^. Over the past two decades, the prevalence of ASD has notably risen: in 2018, approximately 1 in 44 US children were diagnosed with ASD by the age of 8 years ^4^. Early diagnosis can greatly improve a child’s development ^5,6^ and help them realize their full potential, so they can access support and services as soon as possible. Early intensive interventions significantly improve behavioural and social outcomes for children with ASD, including cognitive ability, adaptive behaviour, socialization, and motor skills ^7–9^. However, the diagnosis of ASD currently occurs via recognition of symptomology that is non-systematic and imprecise, resulting in missed and delayed diagnoses ^10,11^. Despite the benefit of early identification of ASD, no universal screening programs exist. Prior research has identified a limited number of risk factors associated with ASD (*e.g.*, complications at birth, family history of ASD, born to older parents, etc.), and routinely collected health data during pregnancy and early childhood offer an opportunity for universal screening and early diagnosis, intervention and support.

Routinely captured medical and health administrative data have been used to develop ASD screening algorithms. Rahman *et al.* combined maternal and paternal electronic medical records for 96,138 patients (1,397 ASD cases, 94,741 controls), and showed that models incorporating prescribed medications, parental age, and socioeconomic status to identify ASD achieved AUROCs of 0.69-0.72 ^12^. Chen *et al.* used diagnostic and procedural codes from medical claims data for 38,576 individuals (12,743 ASD cases, 25,833 controls) to identify ASD in children of 18, 24, and 30 months old, with AUROCs between 0.71-0.87 ^13^. However, these studies only considered a limited number of features. Machine learning (ML) has been used to consider a wide range of features for identifying ASD cases, including structural differences in brain magnetic resonance imaging (MRI) ^14–16^, social and behavioural questionnaires ^17–22^, and gene expression profiles ^23,24^. These ML approaches, while promising, are not ethical or feasible when applied across a population. To date, no predictive models for ASD screening have been developed and evaluated for population-level screening.

Deep learning (DL) algorithms are a class of dense artificial neural networks than can identify complex predictive features from vast volumes of data. DL models can mine comprehensive and granular individual-level records within these datasets and identify features that are most strongly associated with the outcome of interest. Transformer models are a new class of DL models that can be trained more efficiently than previous recurrent neural network architectures.^25,26^ By also incorporating an explainable artificial intelligence (XAI) approach when training DL models, model developers and end users (e.g., healthcare practitioners) can gain insight into how various patient characteristics and other factors contribute to model predictions.^27,28^ In this study, we examine the feasibility of using novel DL models and comprehensive, population-based, and routinely collected health data from Ontario to identify young children with elevated risk of developing ASD.

## 2. Methods

We conducted a retrospective, population-based cohort study using data from ICES - an Ontario-based independent, non-profit research institute whose legal status under Ontario’s health information privacy law allows it to collect and analyze health care and demographic data, without consent, for health system evaluation and improvement. The Children’s Hospital of Eastern Ontario’s Research Ethics Board (REB# 22/06PE) and the ICES Privacy Office (ICES# 2023 901 377 000) approved this study. We implemented two distinct ML algorithms, Transformer and Extreme Gradient Boosting (XGBoost),^25,26,29^ using individual-level mother-infant health data to develop and internally validate a predictive model, leveraging XAI methods to identify features associated with developing ASD. An overview of our methodological framework is presented in Figure 1.

**Figure 1.**
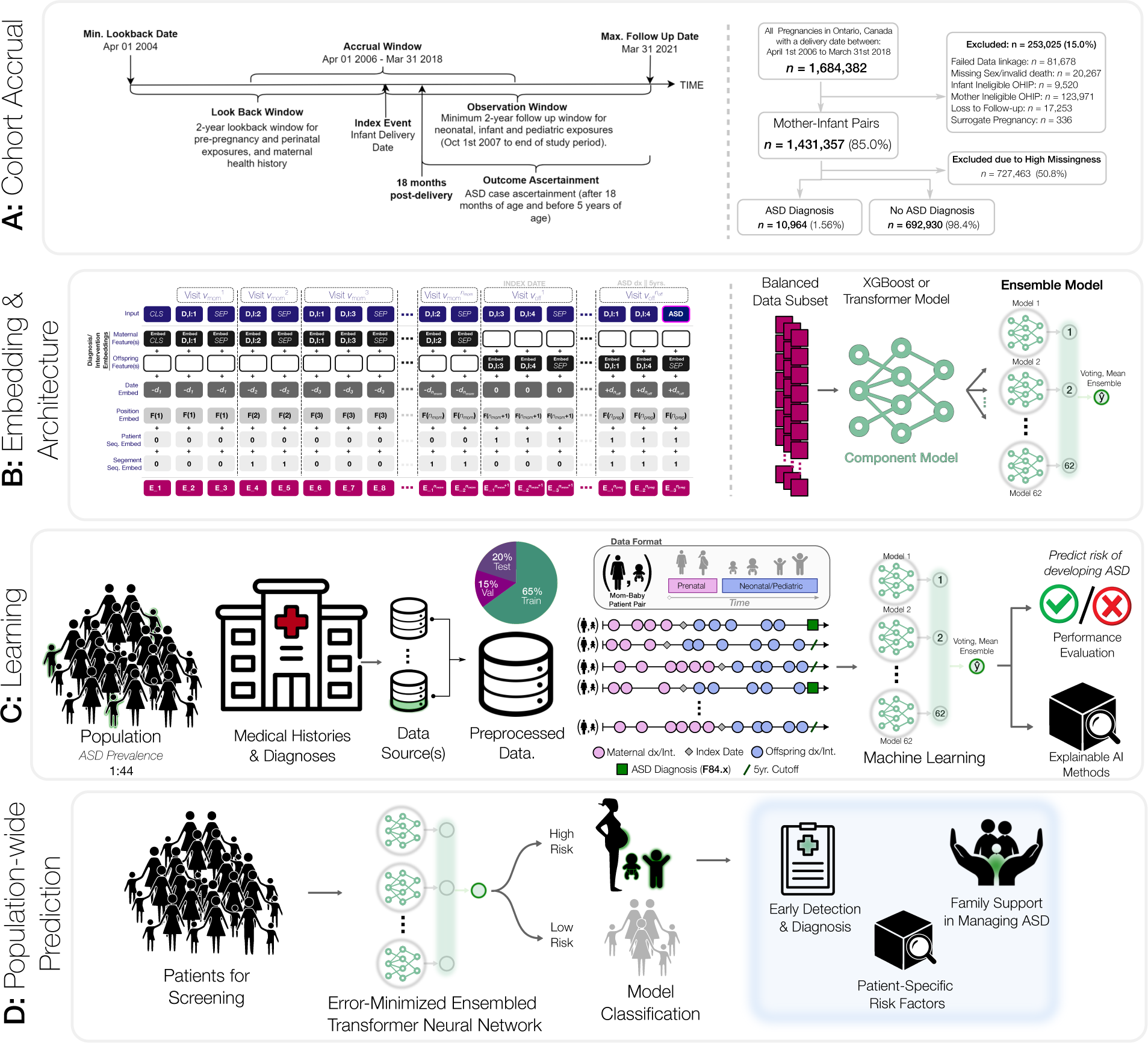
Training and prediction of autism spectrum disorder from mother-offspring health data. **A, Cohort Accrual:** we specify the health data leveraged within our study and their inclusion/exclusion criteria. The maternal look-back window relative to the index is a maximum of 2 years and the offspring observation window is a minimum of 2 years and up to a maximum of 5 years. The outcome of interest is an ASD diagnosis between 18 and 64 months. **B, Embedding & Architecture**: the trained model requires converting real-world clinical data into an embedding – a transformation of categorical disease and intervention codes, timestamps, and related patient data into a lower-dimensional real number continuous space. The transformer model then extracts relevant patterns from the disease history and leverages this latent space to generate ASD risk predictions. To make use of all available data, we trained 62 individual component models and combined their predictions within a large-scale voting ensemble model that outputs a final high-confidence prediction. **C, Learning**: the general ML framework begins by portioning the mother & offspring medical histories into a training set, a validation set, and a test set. The data sources are numerous linked repositories aggregated by mother-offspring ID and preprocessed into both time-series and static one-hot encoded representations for subsequent machine learning algorithm development. The training and validation datasets are used to train the models and minimize the prediction error. **D, Population-wide Prediction:** we evaluate the final model’s prediction performance on the independently held-out test set to quantify its ability to generalize to unseen cases. This final model is used to discriminate between patients at higher and lower risk of developing ASD and this risk model can be leveraged as part of a population screening program.

### 2.1 Data Acquisition and Outcome Definition

This study combines both maternal and offspring characteristics for ASD prediction. Maternal characteristics and medical information prior to and during pregnancy were considered with a look-back period of two years from the offspring’s date of birth (Figure 1C), A follow-up period was applied to collect offspring characteristics, beginning at birth and continuing for five years, until ASD was diagnosed, or until the last available data entry in ICES - whichever occurred first. All datasets were linked using unique encoded identifiers and analyzed at ICES. Our cohort was derived from Better Outcomes Registry & Network (BORN) Ontario:a provincial prescribed perinatal, newborn and child registry.^1^ The cohort consisted of all live births between April 1^st^, 2006 – March 31^st^, 2018, with mother and offspring information linked through the MOMBABY dataset at ICES. This cohort was then linked to additional datasets: Newborn Screening Ontario (NSO), Prenatal Screening Ontario (PSO), Canadian Institute for Health Information (CIHI)’s Discharge Abstract Database (DAD), and CIHI’s National Ambulatory Care Reporting System (NACRS). NSO and PSO datasets contain screening biomarker values and outcomes, whereas DAD and NACRS consist of International Classification of Diseases (ICD-10) diagnostic codes and Canadian Classification of Health Intervention (CCI) intervention codes assigned during hospital and/or emergency visits and outpatient surgeries. Pairs were excluded from the study cohort using the following criteria: failed linkage to other datasets, invalid death dates, missing offspring sex, mothers or offspring ineligible for the Ontario Health Insurance Plan (OHIP) coverage during the entire study period, offspring with missing follow-up information, and offspring resulting from surrogate pregnancies. Any offspring with missing gestational age or birth weight, or with an ASD diagnosis after 5 years of age were also removed. Next, the cohort was limited to births between 2012-2018 due to high levels of missingness of key biomarkers included in NSO and PSO before 2012. Records with more than 50% missing NSO and PSO data, and those without at least one health contact in the DAD/NACRS datasets were removed (Figure 1A).

The primary outcome of interest was diagnosis of ASD between 18 months to 5 years of age. ASD status was ascertained by a case-finding algorithm previously validated in Ontario health administrative data, which assigns a diagnosis of ASD for all those with at least one F84.x ICD-10 diagnostic code within their records from a hospital discharge, emergency department visit, or outpatient surgery, or the OHIP diagnostic code 299.x a minimum of 3 times in 3 years)^30^. This algorithm was clinically validated and found to have sensitivity of 50.0%, specificity of 99.6%, positive predictive value of 56.6% and a negative predictive value of 99.4%. ^30^

### 2.2 Descrip.ve Analyses

Descriptive analyses were conducted to compare characteristics of mother-offspring pairs with an ASD diagnosis to those without (Table 1). Continuous variables were described using means (SDs) or medians (IQRs). Categorical variables were described using frequencies, percentages and standardized mean differences (SMDs).

**Table 1.**
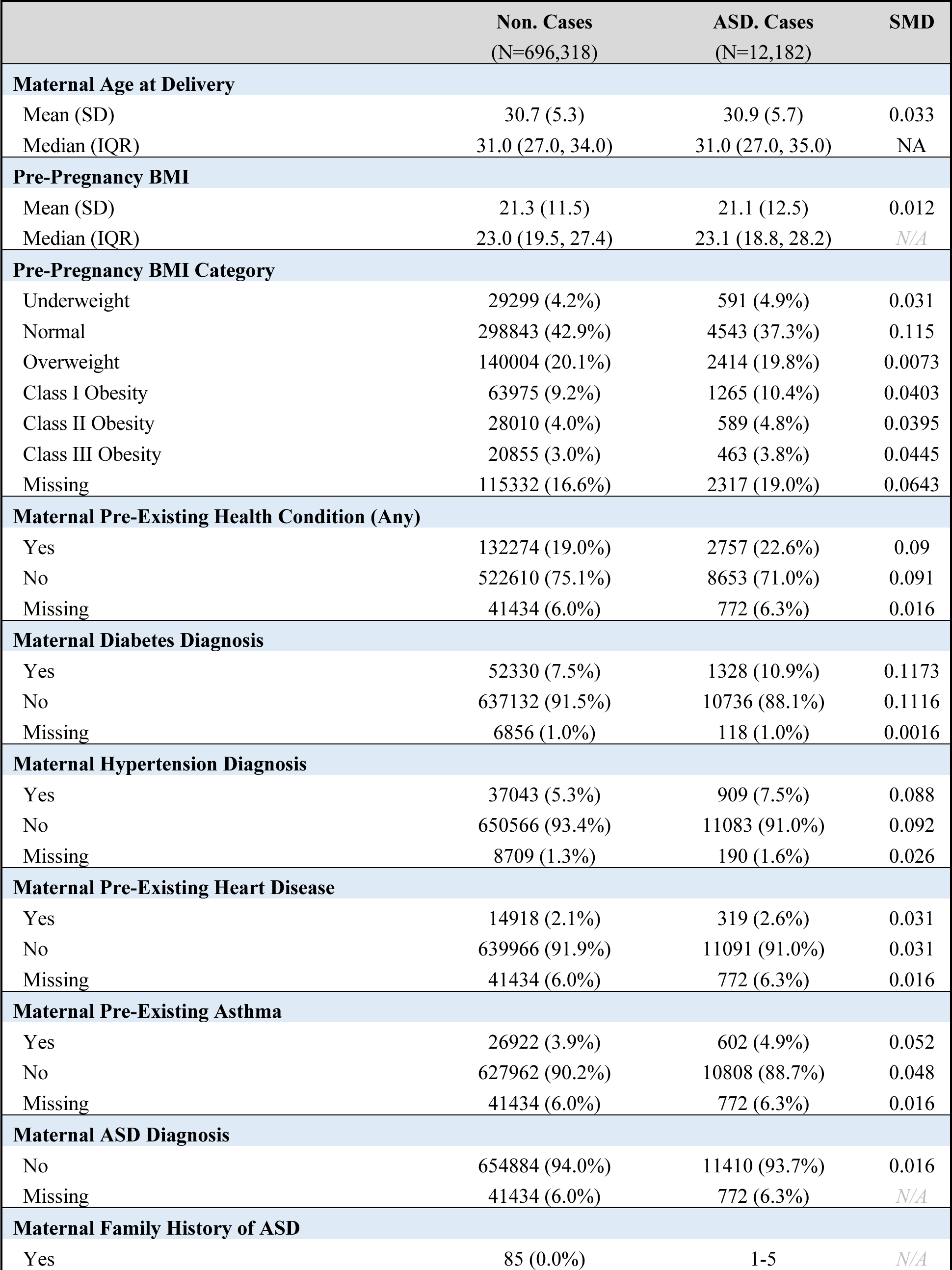
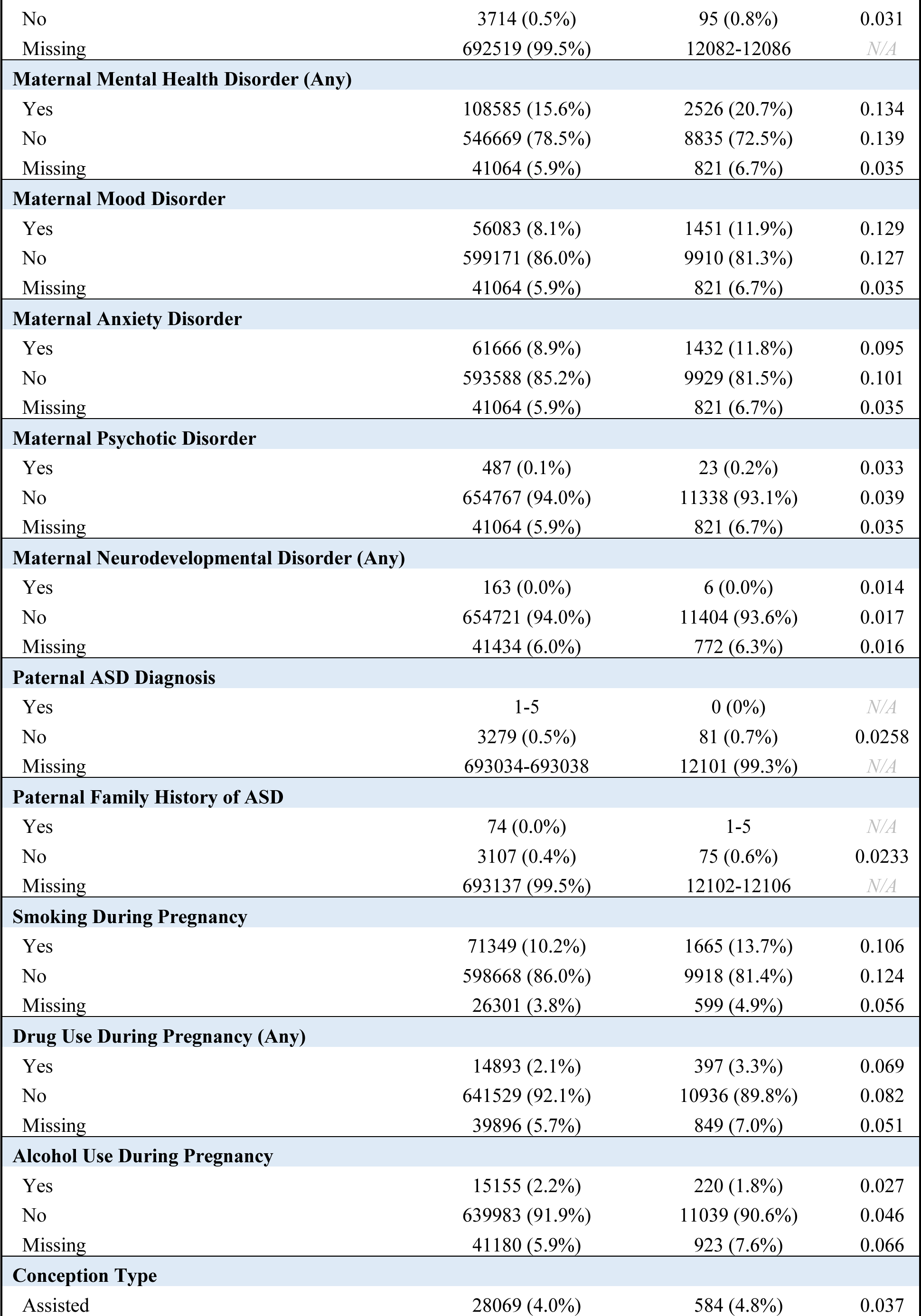
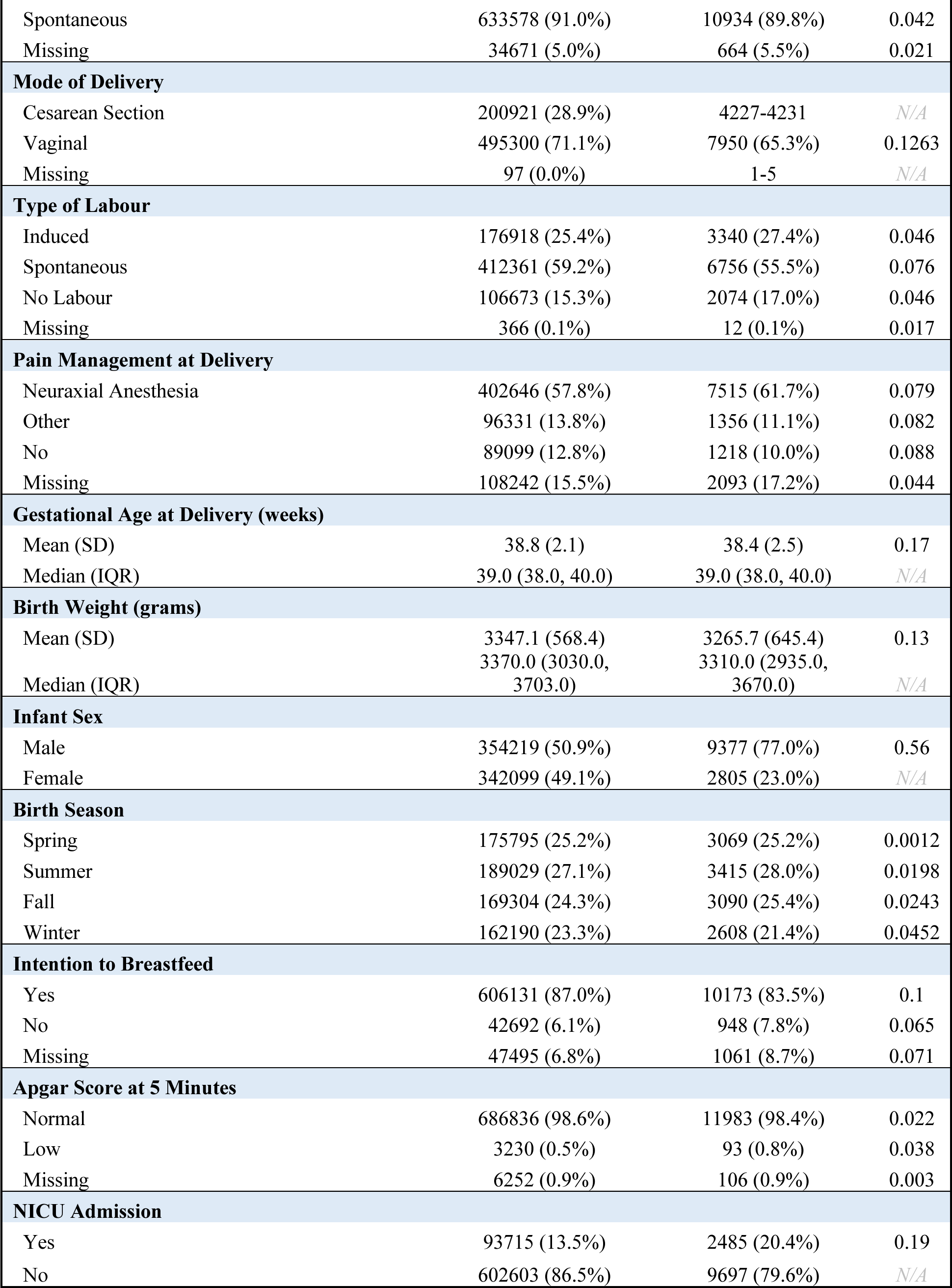
Complete Cohort Descrip4ve Sta4s4cs.

|  | Non. Cases<br>(N=696,318) | ASD. Cases<br>(N=12,182) | SMD |
| --- | --- | --- | --- |
| **Maternal Age at Delivery** |  |  |  |
| Mean (SD) | 30.7 (5.3) | 30.9 (5.7) | 0.033 |
| Median (IQR) | 31.0 (27.0, 34.0) | 31.0 (27.0, 35.0) | NA |
| **Pre-Pregnancy BMI** |  |  |  |
| Mean (SD) | 21.3 (11.5) | 21.1 (12.5) | 0.012 |
| Median (IQR) | 23.0 (19.5, 27.4) | 23.1 (18.8, 28.2) | N/A |
| **Pre-Pregnancy BMI Category** |  |  |  |
| Underweight | 29299 (4.2%) | 591 (4.9%) | 0.031 |
| Normal | 298843 (42.9%) | 4543 (37.3%) | 0.115 |
| Overweight | 140004 (20.1%) | 2414 (19.8%) | 0.0073 |
| Class I Obesity | 63975 (9.2%) | 1265 (10.4%) | 0.0403 |
| Class II Obesity | 28010 (4.0%) | 589 (4.8%) | 0.0395 |
| Class III Obesity | 20855 (3.0%) | 463 (3.8%) | 0.0445 |
| Missing | 115332 (16.6%) | 2317 (19.0%) | 0.0643 |
| **Maternal Pre-Existing Health Condition (Any)** |  |  |  |
| Yes | 132274 (19.0%) | 2757 (22.6%) | 0.09 |
| No | 522610 (75.1%) | 8653 (71.0%) | 0.091 |
| Missing | 41434 (6.0%) | 772 (6.3%) | 0.016 |
| **Maternal Diabetes Diagnosis** |  |  |  |
| Yes | 52330 (7.5%) | 1328 (10.9%) | 0.1173 |
| No | 637132 (91.5%) | 10736 (88.1%) | 0.1116 |
| Missing | 6856 (1.0%) | 118 (1.0%) | 0.0016 |
| **Maternal Hypertension Diagnosis** |  |  |  |
| Yes | 37043 (5.3%) | 909 (7.5%) | 0.088 |
| No | 650566 (93.4%) | 11083 (91.0%) | 0.092 |
| Missing | 8709 (1.3%) | 190 (1.6%) | 0.026 |
| **Maternal Pre-Existing Heart Disease** |  |  |  |
| Yes | 14918 (2.1%) | 319 (2.6%) | 0.031 |
| No | 639966 (91.9%) | 11091 (91.0%) | 0.031 |
| Missing | 41434 (6.0%) | 772 (6.3%) | 0.016 |
| **Maternal Pre-Existing Asthma** |  |  |  |
| Yes | 26922 (3.9%) | 602 (4.9%) | 0.052 |
| No | 627962 (90.2%) | 10808 (88.7%) | 0.048 |
| Missing | 41434 (6.0%) | 772 (6.3%) | 0.016 |
| **Maternal ASD Diagnosis** |  |  |  |
| No | 654884 (94.0%) | 11410 (93.7%) | 0.016 |
| Missing | 41434 (6.0%) | 772 (6.3%) | N/A |
| **Maternal Family History of ASD** |  |  |  |
| Yes | 85 (0.0%) | 1-5 | N/A |
| No | 3714 (0.5%) | 95 (0.8%) | 0.031 |
| Missing | 692519 (99.5%) | 12082-12086 | N/A |
| **Maternal Mental Health Disorder (Any)** |  |  |  |
| Yes | 108585 (15.6%) | 2526 (20.7%) | 0.134 |
| No | 546669 (78.5%) | 8835 (72.5%) | 0.139 |
| Missing | 41064 (5.9%) | 821 (6.7%) | 0.035 |
| **Maternal Mood Disorder** |  |  |  |
| Yes | 56083 (8.1%) | 1451 (11.9%) | 0.129 |
| No | 599171 (86.0%) | 9910 (81.3%) | 0.127 |
| Missing | 41064 (5.9%) | 821 (6.7%) | 0.035 |
| **Maternal Anxiety Disorder** |  |  |  |
| Yes | 61666 (8.9%) | 1432 (11.8%) | 0.095 |
| No | 593588 (85.2%) | 9929 (81.5%) | 0.101 |
| Missing | 41064 (5.9%) | 821 (6.7%) | 0.035 |
| **Maternal Psychotic Disorder** |  |  |  |
| Yes | 487 (0.1%) | 23 (0.2%) | 0.033 |
| No | 654767 (94.0%) | 11338 (93.1%) | 0.039 |
| Missing | 41064 (5.9%) | 821 (6.7%) | 0.035 |
| **Maternal Neurodevelopmental Disorder (Any)** |  |  |  |
| Yes | 163 (0.0%) | 6 (0.0%) | 0.014 |
| No | 654721 (94.0%) | 11404 (93.6%) | 0.017 |
| Missing | 41434 (6.0%) | 772 (6.3%) | 0.016 |
| **Paternal ASD Diagnosis** |  |  |  |
| Yes | 1-5 | 0 (0%) | N/A |
| No | 3279 (0.5%) | 81 (0.7%) | 0.0258 |
| Missing | 693034-693038 | 12101 (99.3%) | N/A |
| **Paternal Family History of ASD** |  |  |  |
| Yes | 74 (0.0%) | 1-5 | N/A |
| No | 3107 (0.4%) | 75 (0.6%) | 0.0233 |
| Missing | 693137 (99.5%) | 12102-12106 | N/A |
| **Smoking During Pregnancy** |  |  |  |
| Yes | 71349 (10.2%) | 1665 (13.7%) | 0.106 |
| No | 598668 (86.0%) | 9918 (81.4%) | 0.124 |
| Missing | 26301 (3.8%) | 599 (4.9%) | 0.056 |
| **Drug Use During Pregnancy (Any)** |  |  |  |
| Yes | 14893 (2.1%) | 397 (3.3%) | 0.069 |
| No | 641529 (92.1%) | 10936 (89.8%) | 0.082 |
| Missing | 39896 (5.7%) | 849 (7.0%) | 0.051 |
| **Alcohol Use During Pregnancy** |  |  |  |
| Yes | 15155 (2.2%) | 220 (1.8%) | 0.027 |
| No | 639983 (91.9%) | 11039 (90.6%) | 0.046 |
| Missing | 41180 (5.9%) | 923 (7.6%) | 0.066 |
| **Conception Type** |  |  |  |
| Assisted | 28069 (4.0%) | 584 (4.8%) | 0.037 |
| Spontaneous | 633578 (91.0%) | 10934 (89.8%) | 0.042 |
| Missing | 34671 (5.0%) | 664 (5.5%) | 0.021 |
| **Mode of Delivery** |  |  |  |
| Cesarean Section | 200921 (28.9%) | 4227-4231 | N/A |
| Vaginal | 495300 (71.1%) | 7950 (65.3%) | 0.1263 |
| Missing | 97 (0.0%) | 1-5 | N/A |
| **Type of Labour** |  |  |  |
| Induced | 176918 (25.4%) | 3340 (27.4%) | 0.046 |
| Spontaneous | 412361 (59.2%) | 6756 (55.5%) | 0.076 |
| No Labour | 106673 (15.3%) | 2074 (17.0%) | 0.046 |
| Missing | 366 (0.1%) | 12 (0.1%) | 0.017 |
| **Pain Management at Delivery** |  |  |  |
| Neuraxial Anesthesia | 402646 (57.8%) | 7515 (61.7%) | 0.079 |
| Other | 96331 (13.8%) | 1356 (11.1%) | 0.082 |
| No | 89099 (12.8%) | 1218 (10.0%) | 0.088 |
| Missing | 108242 (15.5%) | 2093 (17.2%) | 0.044 |
| **Gestational Age at Delivery (weeks)** |  |  |  |
| Mean (SD) | 38.8 (2.1) | 38.4 (2.5) | 0.17 |
| Median (IQR) | 39.0 (38.0, 40.0) | 39.0 (38.0, 40.0) | N/A |
| **Birth Weight (grams)** |  |  |  |
| Mean (SD) | 3347.1 (568.4) | 3265.7 (645.4) | 0.13 |
| Median (IQR) | 3370.0 (3030.0, 3703.0) | 3310.0 (2935.0, 3670.0) | N/A |
| **Infant Sex** |  |  |  |
| Male | 354219 (50.9%) | 9377 (77.0%) | 0.56 |
| Female | 342099 (49.1%) | 2805 (23.0%) | N/A |
| **Birth Season** |  |  |  |
| Spring | 175795 (25.2%) | 3069 (25.2%) | 0.0012 |
| Summer | 189029 (27.1%) | 3415 (28.0%) | 0.0198 |
| Fall | 169304 (24.3%) | 3090 (25.4%) | 0.0243 |
| Winter | 162190 (23.3%) | 2608 (21.4%) | 0.0452 |
| **Intention to Breastfeed** |  |  |  |
| Yes | 606131 (87.0%) | 10173 (83.5%) | 0.1 |
| No | 42692 (6.1%) | 948 (7.8%) | 0.065 |
| Missing | 47495 (6.8%) | 1061 (8.7%) | 0.071 |
| **Apgar Score at 5 Minutes** |  |  |  |
| Normal | 686836 (98.6%) | 11983 (98.4%) | 0.022 |
| Low | 3230 (0.5%) | 93 (0.8%) | 0.038 |
| Missing | 6252 (0.9%) | 106 (0.9%) | 0.003 |
| **NICU Admission** |  |  |  |
| Yes | 93715 (13.5%) | 2485 (20.4%) | 0.19 |
| No | 602603 (86.5%) | 9697 (79.6%) | N/A |

### 2.3 Data Preprocessing

Due to the different strategies required for processing temporal and static data from multiple sources, datasets were preprocessed separately according to distinct protocols (Figure 1B, details in Online Supplement).

### 2.4 Machine Learning

We designed and evaluated two ML algorithms for the prediction of ASD: BEHRT, a transformer developed for the analysis of electronic health records (EHR) ^31^, and XGBoost a boosted tree-based ML model ^32^. BEHRT is a time-series model that analyzes temporal sequences of hospital visits and matching sequences comprising the date of the visit and the patient admitted. These sequences are embedded into latent representations and combined as the input to transformer attention layers, with a final linear layer for prediction of ASD (Figure 1B). To compare performance to that of a non-DL baseline method, we transformed temporal variables into static representation and predicted ASD with XGBoost. We pretrained BEHRT with masked-language-modelling and used the architecture from the original publication that delivered the best performance ^31^. For XGBoost, we applied our previous work, and ran large-scale hyperparameter tuning experiments leveraging high-performance computing infrastructure ^33^. See Supplementary Appendix for complete details.

The data were divided into training, validation, and test partitions with a respective 65:15:20% split, stratifying both by outcome (ASD vs. non-ASD) and maternal identifier (maternal/parental features only appear within independent sets) (Figure 1C). This two-part stratification ensures equal representation of ASD and prevents data leakage of the same mother appearing in both data partitions. Any transformations applied to numeric variables were first performed on training data; the same parameters were then used to transform the validation and test data.

Training individual models with balanced training data (downsampling the majority class to 1:1) would result in the loss of 469,051 of non-ASD cases, limiting the generalizability of our findings. We used large-scale ensemble of component models for our final model, where each model was trained with the same 7,624 of ASD cases and a different subset of non-ASD controls (sampled without replacement), ultimately leveraging all available data. The testing data was then evaluated on each model, and majority voting was used for final ASD prediction. This approach was applied to both the BEHRT and XGBoost model architectures.

### 2.5 Evaluation Metrics

We assessed prediction performance, for all models trained and evaluated, by measuring sensitivity (*i.e.,* true positive rate or recall), specificity (*i.e.,* true negative rate), and positive predictive value (PPV or precision) at different risk thresholds (Figure 2). As depicted in Figure 1D, an error-minimized model could be leveraged in the future as a component of a population screening program. The model’s overall ability to discriminate was determined using the area under the receiver operating characteristic curve (AUROC). Given the extreme class imbalance, we additionally report the F1 score (defined as the harmonic mean of PPV and sensitivity) and the area under the precision-recall curve (AUPRC). Finally, we reported the specificity and PPV value where the predicted probability cutoff yields a sensitivity of 50%. Cumulative gain curves were also created to express that our model can be used to identify an enriched pool of high-risk children.

**Figure 2.**
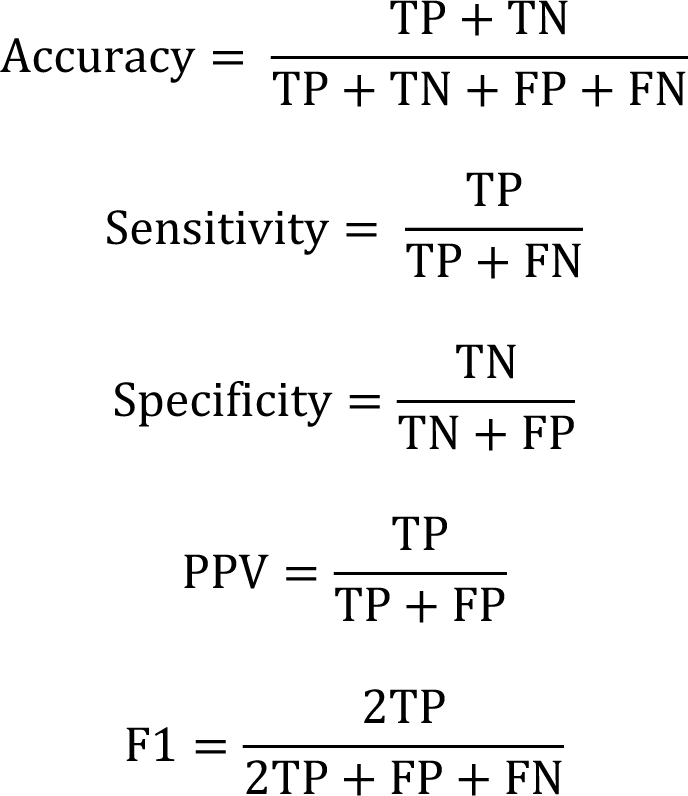
Performance metrics used for all models. TP: true positives (number of instances correctly predicted to be positive); TN; true negatives (number of instances correctly predicted to be negative), FP: false positives (number of instances incorrectly predicted to be positive); FN false negatives (number of instances incorrectly predicted to be negatives).

### 2.6 Algorithm Explainability

To identify the most predictive features for ASD risk, we used game-theoretic SHapley Additive exPlanations (SHAP) analysis ^34^ to query the trained models and obtain an indication of how significant each factor is in determining the final ASD prediction ^35^. SHAP analysis generates many prediction experiments that vary ‘coalitions’ (or feature combinations) to compare the impact of variable inclusion/exclusion against the other features to quantitatively assess the average impact of a given feature on the overall model ^34^.

## 3. Results

The final study cohort included 703,894 mother-infant pairs, from deliveries between 2012 and 2018, including 10,964 ASD cases (1.56%) (Figure 1A). We observed imbalance between outcome groups in several maternal and infant characteristics (Table 1). Compared to children without ASD, more children with ASD were male, were delivered by caesarean section, were admitted to the NICU, had a lower mean birth weight, and were younger gestational age at delivery. In addition, pre-existing diagnoses of maternal mental health disorders or diabetes were more prevalent in mothers to children with ASD, compared to those to children without ASD. Reported smoking during pregnancy was also more prevalent in mothers of children with ASD.

Table 2 lists the results of the hyperparameter tuning experiments and final model performance. Resampling experiments revealed that the highest performance (using high sensitivity as the objective) was achieved from downsampled balanced ASD and non-ASD cases during training, motivating the development of a large-scale ensemble model. The final best-performing ensemble model achieved an AUROC of 69.6%, a sensitivity of 70.9%, a specificity of 56.9% and a positive predictive value of 2.4%.

**Table 2.**
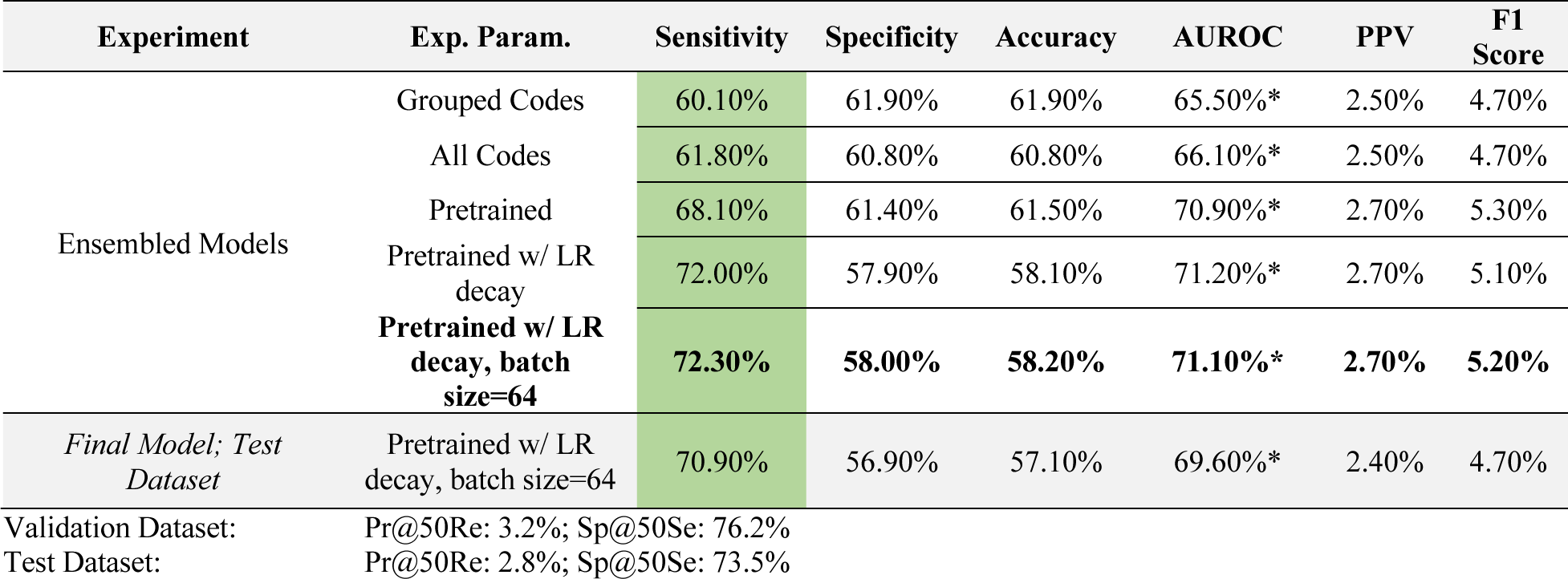
Hyperparameter tuning Transformer models on the valida4on dataset & evalua4ng generalizability on the test datasets. Final model selected by highest sensi2vity on the valida2on dataset (bold). * indicates the *mean* metric of the ensembled models rather than the *vo(ng ensemble* metric.

| Experiment | Exp. Param. | Sensitivity | Specificity | Accuracy | AUROC | PPV | F1 Score |
| --- | --- | --- | --- | --- | --- | --- | --- |
| Ensembled Models | Grouped Codes | 60.10% | 61.90% | 61.90% | 65.50%* | 2.50% | 4.70% |
|  | All Codes | 61.80% | 60.80% | 60.80% | 66.10%* | 2.50% | 4.70% |
|  | Pretrained | 68.10% | 61.40% | 61.50% | 70.90%* | 2.70% | 5.30% |
|  | Pretrained w/ LR decay | 72.00% | 57.90% | 58.10% | 71.20%* | 2.70% | 5.10% |
|  | **Pretrained w/ LR decay, batch size=64** | **72.30%** | **58.00%** | **58.20%** | **71.10%\*** | **2.70%** | **5.20%** |
| *Final Model; Test Dataset* | Pretrained w/ LR decay, batch size=64 | 70.90% | 56.90% | 57.10% | 69.60%* | 2.40% | 4.70% |
| Validation Dataset: | Pr@50Re: 3.2%; Sp@50Se: 76.2% |  |  |  |  |  |  |
| Test Dataset: | Pr@50Re: 2.8%; Sp@50Se: 73.5% |  |  |  |  |  |  |

The receiver operator characteristic (ROC) curves for the voting and mean ensemble Transformer model for validation and test datasets are illustrated in Figure 3. Consistent performance curves across the validation and test datasets indicates our model did not overfit the training data and generalizes well to unseen data. From the cumulative gain plots in (Figure 4), we note that the top-5% of the model’s prediction contain approximately 15% of all true cases and the top-10% of model predictions contain 25% of all true cases, suggesting that our model can be used to identify an enriched pool of high-risk cases.

**Figure 3.**
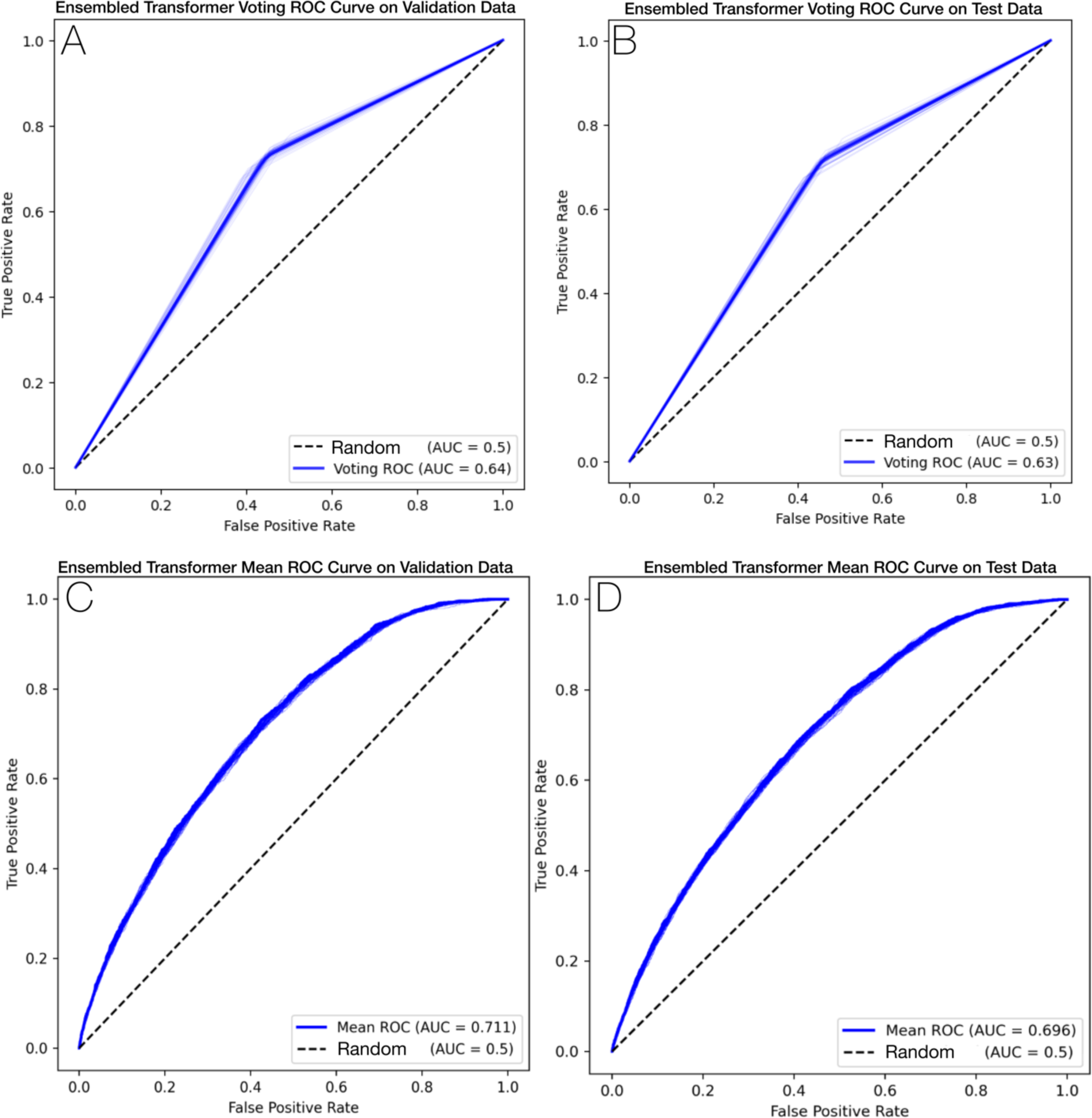
ROC curve summarizing performance of the final ensembled transformer model on the validation and test datasets. All *n* = 62 individual component model curves are plotted in light blue, overlaid but the voting (A,B) and mean (C, D) ROC curves summarizing the overall performance with respect to the validation (A, C) and test (B, D) datasets.

**Figure 4.**
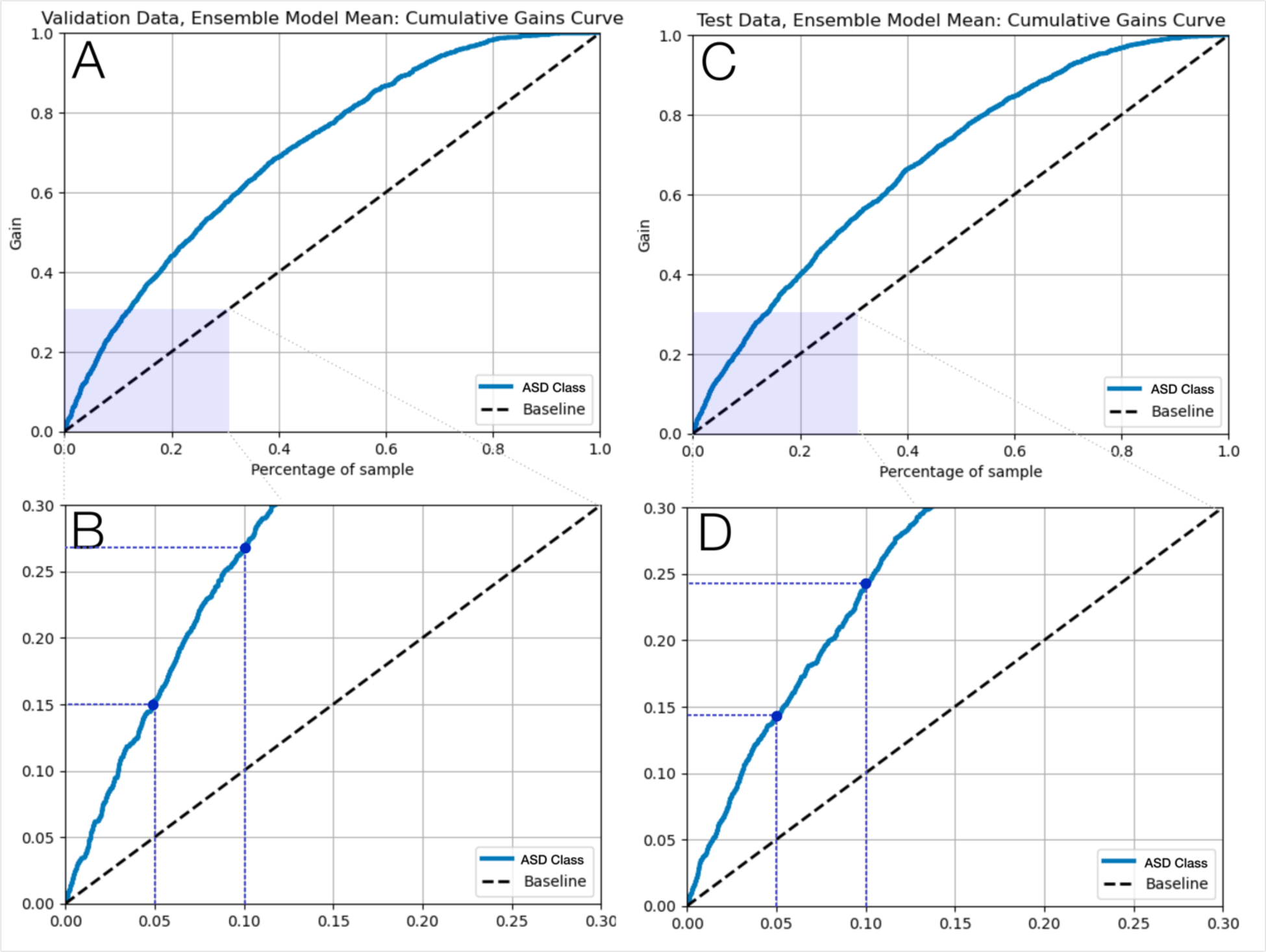
Ensemble transformer mean model cumulative gain curves.

An illustration of the top-ranking features identified via SHAP analysis across the three independent datasets is presented in Figure 5, where a positive SHAP value suggests that the factor increases the predicted probability of ASD while a negative value suggests that the feature decreases the predicted probability of ASD. Interestingly, a mixture of BORN-BIS, NSO, ICD-10, and DAD/NACRS features rank among the top-20 of each set with a general consistency.

**Figure 5.**
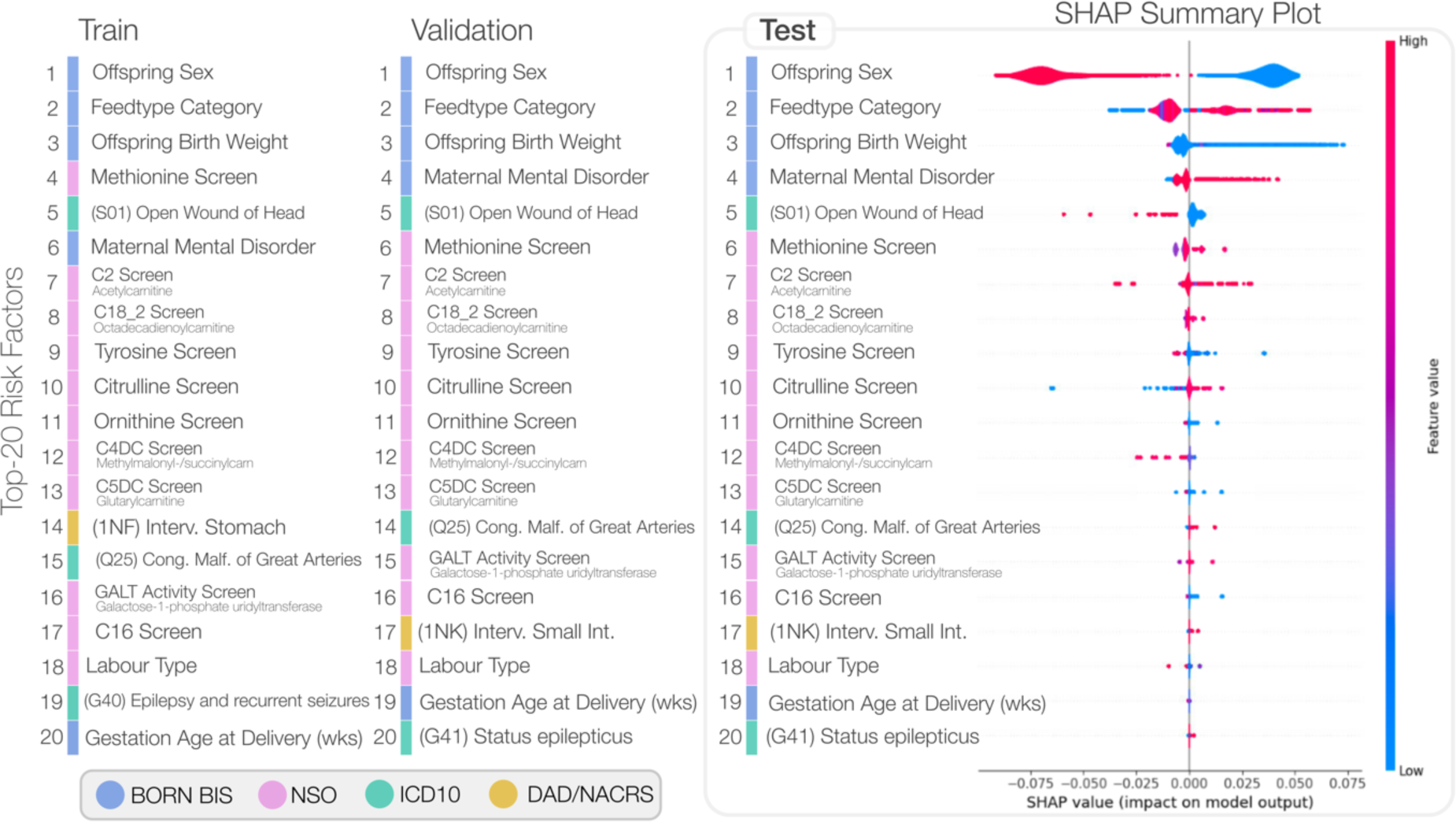
Summary of the top-ranking risk factors determined using SHAP analysis across the three independent datasets. The right-most SHAP summary plot depicts the violin plot distribution for each factor. The possible feature values for each variable are tabulated in Supplementary Table S3; individual KDE plots comparing ASD and controls are illustrated in Supplementary Figure S2.

## 4. Discussion

In this retrospective, population-based cohort study from Ontario, Canada, we designed and implemented ML models to predict ASD in our final study cohort of 703,894 mother-offspring pairs. The best-performing ensemble of Transformer models achieved an AUROC of 69.6% for predicting ASD diagnosis, a sensitivity of 70.9%, a specificity of 56.9%; results that are consistent with similar studies such as the work of Betts *et al.* ^36^. We applied ML best practices for training a predictor of ASD: comprehensive evaluation metrics, stratified train-test splits, and robust models that address class imbalance. We demonstrated model generalizability given that both Figure 3 and Figure 4 illustrate similar performance across validation and test datasets and Figure 5 shows that the most impactful model features and ordering are also consistent across train, validation, and test datasets.

The results of this work demonstrate feasibility and potential to identify young children with increased likelihood of developing ASD using a ML model applied to population-based and routinely collected data. The models presented within this work also have demonstrated face validity given that top-ranking predictive features (Figure 3) include known ASD risk factors (e.g., male sex, low birth weight). By incorporating large-scale and heterogenous datasets, these models and XAI features provide testable hypotheses for future work. Our models highlight a number of newborn screening factors that, following additional investigation, could be incorporated within an early life universal ASD screening program.

To our knowledge, the only other studies to have used EMR ICD codes and a similar ML methodology were conducted by Betts *et al.* and Bishop-Fitzpatrick *et al.* ^36,37^. Our work reveals significant advances over these studies. The Betts *et al.* dataset ranged from 2003-2005 with approximately 260,000 offspring ^37^, whereas our work includes ∼700,000 offspring and spans 2012-2018. While earlier models utilized Logistic Regression and XGBoost models ^37^, we apply Transformer-based models that sequentially analyze mother-infant medical histories. Regarding performance, our work achieves a slightly lower AUROC score of 70% compared to the 73% reported by Betts *et al.* ^37^. The fact tha we demonstrate similar performance is significant given that these studies originate from completely different international settings. The complementary use of SHAP analysis as an XAI method within both studies offer promising insights into the candidate factors that may assist healthcare providers in understanding this complex neurodevelopmental condition. Finally, our use of a large-scale ensembling model architecture to address extreme class imbalance typical within healthcare dataset (as well as our approach to extensive hyperparameter tuning) are notable contributions that advance conventional applied healthcare ML methodologies.

One of the limiting factors of all studies using medical claims for prediction of ASD, including ours, is the increased number of hospital/doctor’s visits that ASD patients have compared to normal cases. This may introduce bias into the data, where the ML model begins to make predictions based on the number of visits a child has, as opposed to true ASD risk factors. While this is inherent in any medical claims data, our study mitigates this risk by combining offspring visits with maternal visits and padding/truncating all visits/codes to a length of 200. In addition, we choose to include the entire cohort of non-ASD controls, as opposed to selecting a subset for model training and evaluation. Although we have a similar number of total ASD cases to Chen *et al* ^13^., our model can better generalize to the entire population for screening ASD due to the inclusion of all possible population data. Other limitations include the lack of paternal information, imposing a bias and unequitable focus on maternal factors. Unfortunately, paternal medical data is not reliably collected and should be the subject of future research.

Another important limiting factor of this work originates from the ICES ASD algorithm from which the ASD ground truth labels are acquired. The work of Brooks *et al.* assessed numerous algorithms for the identification of children with ASD in health administrative datasets resulting in the labels leveraged in this work ^30^. Their optimal algorithm achieved a sensitivity of 50.0% (95% CI 40.7–88.7%), specificity of 99.6% (99.4–99.7), PPV of 56.6% (46.8–66.3), and NPV 99.4% (99.3–99.6) ^30^. Given the performance of this algorithm in establishing the ground truth for ASD in our own study, we cannot expect the models produced herein to exceed this level of performance.

Bias must be considered before deploying any clinical decision-support tool that incorporates ML for early detection. Biases can originate in the data used, the algorithm, or a combination of both. For instance, in our study focusing on childhood ASD, the manifestation of the condition prevalence differs significantly between males and females (sex assignment at birth). This discrepancy leads to a lower rate of diagnosis and, consequently, a reduction in available treatment in female patients. If not fully considered and transparently understood, models like ours could unintentionally exacerbate this gender disparity. Our model may result in increased likelihood of misclassification among females, erroneously classifying females with ASD as controls in the sample data. Additionally, the skewed representation of male patients in the training data may unintentionally cause the algorithm to optimize for male-related indicators, thereby enhancing prediction accuracy for males while diminishing it for females. To prevent such discriminatory outcomes, it is imperative to thoroughly evaluate model performance across different patient subgroups and implement measures to mitigate these biases.

Future work will seek to improve performance and identify new datasets for predicting ASD via our framework. Universal screens must limit the number of features considered by the model to those most reliable and impactful to consistently distinguishing ASD in young children. Thus future will work will also apply an ablation-like approach to ensure that we develop and implement a model for which the data is reliably collected, and the clinical relevance of the features are well-understood. Ultimately, such an AI-based ASD screen will have the potential for deployment and use as a clinical decision support system, as depicted in Figure 1D. Additionally, the methodology and framework developed within this study could be applied to other complex neurodevelopmental conditions, towards a multi-condition screening framework.

## 5. Conclusion

This study demonstrates the feasibility of applying ML models to population-based and routinely collected health information to systematically identify young children who are likely to develop ASD. Evaluated on a fully independent dataset representative of a general population sample, our model’s reported sensitivity of 70.9%, specificity of 56.9%, and AUROC of 69.6% suggest that our ensemble transformer model is a promising candidate for population-based ASD screening. Early identification through this method could facilitate comprehensive and timely assessment for ASD, ensuring prompt diagnosis and faster access to resources, support, or therapy.

## Data Availability

All data produced in the present study are available upon reasonable request to ICES.

## 7. Acknowledgments

This project is supported by an anonymous donation to develop the CHEO Precision Child and Youth Mental Health Initiative. This study was additionally supported by ICES, which is funded in part by an annual grant from the Ontario Ministry of Health (MOH). This study was based on data compiled by ICES and on data and/or information compiled and provided by CIHI. However, the analyses, conclusions, opinions, and statements expressed herein are those of the author(s), and not necessarily those of ICES or CIHI. We thank Arya Rahgozar, Ottawa Hospital Research Institute, for reviewing and providing feedback on an early draft of this manuscript. We thank Carolina Lavin-Venegas, BORN Ontario and CHEO Research Institute, for her assistance with manuscript submission.

## Supplementary Appendix

### Supplementary Methods

#### 1. Temporal Data Preprocessing

The diagnostic and intervention code datasets (DAD, NACRS) were acquired from ICES as large data frames, with each row representing a single hospital/emergency visit. Each diagnosis and intervention code assigned to a patient during a specific visit was separated into an individual column. All infant visits (for ASD cases and non-cases) occurring after 5 years of age were censored, and for ASD cases, the visit with the initial ASD diagnosis was censored, in addition to all visits following the diagnosis date. Across all remaining visits, there were a total of 17,142 unique diagnosis and intervention codes. In preparation for input to BEHRT, DAD and NACRS information for a single patient were concatenated to form a sequence of temporal medical visits. Following terminology defined in BEHRT (Figure 1D), each patient’s (*i.e.*, mother or offspring) medical history can be defined as a series of visits, *V_p_ =* {*v_p_*^1^*, v_p_*^2^*, v_p_*^3^*,… v_p_^np^*}, where each patient, *p*, has a total of *n_p_* visits. Each individual visit, *v_p_^j^*, is defined by *m* number of diagnosis and/or intervention codes *c* assigned to each patient *p* for visit *j*, given by *v_p_^j^* = {*c_p_*^1^*, c_p_*^2^*, c_p_*^3^*…c_p_^mj^*}. For each patient, visits were temporally ordered and concatenated together, with ‘*SEP*’ added between visits to distinguish one from another. Further, we then concatenated mother visit *V_mom_* sequences with their respective offspring visit sequences *V_off_* to represent visits throughout the entire study timeline, beginning two years prior to birth and ending with the last infant hospital visit before 5 years of age. Similar to BEHRT, we add ‘*CLS*’ at the beginning of the entire sequence. This resulted in a final sequence of *V_preg_ =* {*CLS*, *v_mom_*^1^*, SEP*, *…, v_mom_^nmom^, SEP, v_off_*^1^*, SEP, …, v_off_ ^noff^*}.

To consider temporal information and distinguish between mother and offspring codes, we developed two additional sequences which can be simultaneously input to BEHRT with the code sequence previously described. First, the timeframe between diagnosis and/or intervention codes and their proximity to the birthdate of the offspring may impact ASD diagnosis. Thus, the date of each visit defined in *V_preg_* relative to the birth of the infant is used to create a sequence of dates, *D_p_ =* {*d_p_*^1^*, d_p_*^2^*, d_p_*^3^*,… d_p_^np^* }, or *D_preg_ =* {*d_mom_*^1^, *d_mom_*^1^*, d_mom_*^1^*,…, d_mom_^nmom^, d_mom_^nmom^, d_off_*^1^*, d_off_*^1^*,…, d_off_ ^noff^*} to match the *V_preg_* sequence defined above. To match the length of the visit sequence, the date is repeated for each code assigned during that visit, and all ‘*SEP*’ and ‘*CLS*’ values. Therefore, for a single visit, all dates will be identical. Next, while the date sequence implicitly defines if a code is assigned to the mother or offspring (all negative dates correspond to the mother, and all positive dates correspond to the child), we also explicitly assign each visit to a patient. The same diagnosis or intervention code may have higher significance for the mother or offspring, and as such, the patient should be explicitly defined within the input to BEHRT. This sequence consists of single integers to separate one patient from another. In our case, we assign 0 to all visits corresponding to the mother (up to the index date) and switches to 1 for infant visits. As with the date sequence, the values are repeated for all diagnosis and intervention codes, as well as ‘*SEP*’ and ‘*CLS*’ values. Therefore, *P_preg_ =* {0, 0, 0, …, 0, 0, 1, 1, …, 1}.

To compare our modified BEHRT time series-based model to the state-of-the-art XGBoost ML architecture, we transformed DAD and NACRS data into static data variables. Rather than one-hot encoding 17,142 individual diagnosis and intervention codes, only a subset of codes were used to define specific health conditions. Specifically, we created variables to define the presence of: maternal diabetes, hypertension, heart disease, asthma, ASD, attention-deficit/hyperactivity disorder, mental health disorder, mood disorder, anxiety disorder, psychotic disorder, or neurodevelopmental disorder diagnosis; smoking, alcohol, or drug use during pregnancy; conception type, mode of delivery, labour type, intervention type during delivery; and/or NICU admission. These variables were also augmented with data from BORN for full completeness. ICD-10 diagnosis codes, CCI intervention codes, and BORN data used to define each variable can be found in Supplementary Data. In addition to these 19 created variables, we also grouped codes based on their first three characters, for a further 2,669 static DAD and NACRS variables.

#### 2. Sta6c Data Preprocessing

Following the preprocessing of temporal data, we next transformed all static data variables from NSO, PSO, and overall cohort information included in this study. The following transformations were applied to the numeric variables. First, all values were log-transformed; values of zero and under were imputed with half of the lowest non-negative value in the dataset prior to transformation. Next, values were pareto standardized, using the square root of the standard deviation in the denominator. Winsorization was then applied to remove all outliers. Lastly, all missing values were imputed using predictive mean matching using the miceforest package in Python.

Of the 142 available NSO variables, 49 were selected based on low missingness and possible relation to ASD based on clinical judgement, and numeric variables were transformed. We also selected five overall cohort variables: offspring birth season, gender, gestational age, birth weight, and maternal age.

#### 4. Sub-analyses

For PSO, there are numerous different prenatal screening tests that can be administered, depending on multiple factors. For this reason, there is a large variation in the completeness of PSO variables limiting the number of samples available to this analysis. To mitigate this, we originally omitted all PSO data from our experiments and instead performed a PSO sub-analysis once the final model was determined. The sub-analysis cohort was limited to the 301,829 pregnancies that received IPS PSO testing. Five numeric PSO variables were chosen and transformed as described above. In addition, four variables were one-hot encoded to define whether an offspring tested positive for trisomy-21, trisomy-18, open neural tube defect, or any rare disorder. The full list of static data variables included can be found in the accompanying DCP. The results of this PSO sub-analysis are available in Table S2where we note that the inclusion/exclusion of PSO variables does not greatly impact the model performance.

#### 5. Machine Learning Methods

We examined two separate machine learning algorithms for the prediction of ASD: BEHRT, a transformer-based deep learning model ^31^ developed for analysis of Electronic Health Records (EHR), and Extreme Gradient Boosting (XGBoost) (27).

##### 5.a. *BEHRT*

To consider temporal information in the prediction of ASD, we adapted BEHRT ^31^. BEHRT is a modification of BERT, a state-of-the-art transformer model designed for natural language processing tasks. Specifically, BERT analyzes sentences (part of larger documents) and learns relationships between words (and their positions in sentences) to understand the context of the sentences. BEHRT uses this same idea to understand relationships in EHR. Modelling codes as words, individual physician/hospital visits as sentences, and entire EHR histories as documents, BEHRT learns relationships between codes to identify how codes are related, understand the context of hospital visits, and predict future codes assigned to a patient. In addition, BEHRT also encodes the age of a patient at the time of an EHR visit to determine the effect of age on the relationships between codes and the prediction of future EHR visits. We modified BEHRT to encode time relative to offspring birth as opposed to age, and to analyze both mother and offspring diagnosis and intervention codes through our patient sequences *P_preg_*.

Our modified BEHRT takes in five individual sequences to understand mom-offspring information: code, date, patient, positional, and segment sequences. As described previously, code sequences represent all diagnosis and intervention codes assigned to a patient during a specific medical visit. Both mother and infant codes were concatenated together. The date and patient sequences represent the date the codes were assigned, and the patient (i.e., mom vs offspring) they were assigned to. As described in BEHRT, positional sequences are used to identify the position of a visit in a patient’s entire medical history (*e.g.*, first medical visit in study timeframe vs fifth medical visit). Positional sequences were encoded through a popular representation created by Vaswani *et al.* ^38^. Segment sequences explicitly separate one visit from another and alternate between 0 and 1. All date, patient, positional, and segment values are repeated for all codes assigned during a given visit, resulting in five separate, same-length sequences.

The BEHRT architecture is designed as follows. Each individual sequence (*e.g.,* code, date, etc.) is input to an embedding layer, which generates a unique latent representation of the information in the sequence. The embeddings are then added together to create a combined representation of the diagnosis/intervention code, the date it was assigned, who it was assigned to, its position within all visits throughout the study timeline, and which segment it belongs to. This combined representation will be termed a ‘code representation’ for the remainder of this manuscript. The code representation is then used as input to the transformer attention layers. BEHRT uses self-attention mechanisms to simultaneously update latent representations while determining importance of each code representation for the given task. Detailed information regarding attention mechanisms can be found in the original articles.

##### 5.b. XGBoost

The concept of gradient boosting machines, initially proposed by Friedman ^39,40^ serves as the foundation for Extreme Gradient Boosting (XGBoost) models ^32^. Similar to Random Forest models, XGBoost models consist of an ensemble of classification and regression trees (CART). These models leverage systems optimization and fundamental machine learning principles; in essence, they maximize computational capabilities, allowing for scalability, portability, and notable accuracy. The fundamental concept behind boosting involves assigning equal initial weights to each sample and iteratively adjusting these weights ^39^. In each iteration, a training set is constructed based on the sample weights, where samples with higher weights have a greater likelihood of being included. Subsequently, a decision tree is constructed using this training set. Following each training iteration, samples that were misclassified during training receive increased weights. The models are then weighted based on the influence of the current model on decision-making, as each model can only accurately learn a portion of the samples, rendering them “weak” models ^39^. Ultimately, the weighted combination of these weak models forms a robust model with enhanced predictive power.

#### 6. Architecture Design and Hyperparameter Tuning

We pre-trained BEHRT with masked-language-modelling (MLM). MLM takes input sequences and masks a certain number of code representations within an entire patient’s medical history. The model is then trained to predict which codes are masked. By training the model to identify the missing codes based on other codes within a patient’s history, relationships between diagnosis/intervention codes are learned by the model. This provides the model with context for all possible codes across all patients, as opposed to inputting codes with no pre-training (which would effectively be meaningless to the model at the start of training). The learned context may improve performance of predicting ASD by allowing the model to better recognize related codes and how they may be associated with an ASD diagnosis. BEHRT is pre-trained with MLM for 30 epochs with randomly initialized code, date, patient, positional, and sequence embedding weights. We applied the methods outlined in BEHRT ^31^, selecting 12% of codes to be masked, and 1.5% of codes to be randomly replaced with other codes.

In this study, we used the same architecture determined to have best performance in the original BEHRT article ^31^. Specifically, we chose 6 hidden layers with a size of 288, 12 attention heads, an intermediate layer size of 512, a learning rate of 3e-5, and a dropout rate of 0.01. All sequences were truncated to a maximum of 200 codes per pregnancy. After pre-training with MLM, the final layer of the network used for MLM predictions was removed and replaced with a linear layer consisting of a single output to predict ASD vs. non-ASD. Prior to the final prediction layer, all static data variables (*i.e*., the 19 static DAD/NACRS variables, 49 NSO variables, and offspring birth weight, birth season, gestational age, sex, and maternal age) were concatenated with the output from the BEHRT attention layers. The network was then trained for an additional 10-25 epochs. For ASD prediction, a loss function of binary cross-entropy with logits loss function was used with a learning rate of 3e-5.

We focused on hyperparameter tuning related to reducing class imbalance due to the low prevalence of ASD. Specifically, we varied the amount of upsampling of the minority class using PyTorch’s Weighted Random Sampler function. This function randomly samples the training data points with replacement; we set the number of overall samples to the original number of training samples, and used various weighting schemes with higher sampling of the minority class. The random sampling therefore simultaneously upsamples the minority class and downsamples the majority class. We also experimented with equal representation of ASD and non-ASD cases by downsampling non-ASD controls to a total of 7,624.

Finally, for the comparative XGBoost model, following the work of Dick *et al.*, we ran large-scale hyperparameter tuning experiments leveraging high-performance computing infrastructure ^33^. Given the large-scale dataset of complex heterogenous data available within this work, we considered a very large hyperparameter space varying the learning rate, maximum tree depth, and the number of estimators for a total of *n=*6,417 independently trained and evaluated models. To visualize and rank top-performing models, we represent model validation performance metrics as a comprehensive heatmap based on a specific metric of interest. Each of the nine subplots depicts the results keeping the learning rate fixed as we vary the maximum tree depth (x-axis) between [3,18] by increments of 1, the number of estimators (y-axis) between [50,600] by increments of 25. Within each subplot, we highlight the maximum value with a black bounding box and the median value with a white bounding box. All results are normalized to the same colour range where lighter values represent better performing models.

## Supplementary Results

Within this section, we present additional experimental results to complement the findings presented in the main text. Notably, the up-/downsampling experiments for individual Transformer models (non-ensemble) are summarized in Table S1. The high sensitivity value achieved from 1:1 downsampling inspired the n=62 component model ensemble transformer model presented in the main text.

**Table S1.** Hyperparameter tuning Transformer models using up-/downsampling of classes on the valida4on dataset. Experiments indicate that training upon a balanced dataset through downsampling the majority class leads to the highest model sensi2vity.

| Experiment | Exp. Param. | Sensitivity | Specificity | Accuracy | AUROC | PPV | F1 Score |
| --- | --- | --- | --- | --- | --- | --- | --- |
| Upsampling Minority Class | None | 0.00% | 100.00% | 98.40% | 66.90% | 0.00% | 0.00% |
|  | 1:2 | 0.20% | 100.00% | 98.40% | 65.40% | 10.00% | 0.40% |
|  | 1:5 | 4.90% | 98.60% | 97.10% | 65.60% | 5.20% | 5.00% |
|  | 1:50 | 9.80% | 94.40% | 93.00% | 62.80% | 2.70% | 4.20% |
|  | 1:63 (balanced) | 8.00% | 95.90% | 94.50% | 63.30% | 3.00% | 4.30% |
|  | 1:120 | 13.40% | 88.70% | 87.50% | 63.10% | 1.90% | 3.30% |
| Downsampling Majority Class | 1:1 | 86.90% | 30.40% | 31.20% | 64.90% | 2.00% | 3.80% |

Furthermore, Table S2 summarises the sub-analysis results when including/exclusing the PSO variables. Given that this sub-analysis was only applied to a limited dataset (only a small fraction of cases undergo PSO screening), the performance values can only be compared to each other and cannot be compared to other tabulated metrics. We determine that the inclusion of PSO variables does not lead to an increase in a meaningful increase in performance metrics.

**Table S2.** Limited dataset sub-analysis of component performance with and without PSO data.

| Experiment | Exp. Param. | Sensitivity | Specificity | Accuracy | AUROC | PPV | F1 Score |
| --- | --- | --- | --- | --- | --- | --- | --- |
| PSO Variable Subanalysis | With PSO | 74.40% | 52.70% | 53.00% | 69.80% | 2.70% | 5.10% |
|  | Without PSO | 73.20% | 54.70% | 55.00% | 70.50% | 2.70% | 5.30% |

## Hyperparameter Tuning of the XGBoost Model(s)

Following from the work of Dick *et al* ^33^, we performed large-scale hyperparameter tuning experiments to train thousands of XGBoost models. Three hyperparameters defining the size and complexity of the XGBoost model were selected and visually summarises by validation dataset recall in Figure S1: the learning rate (LR; individual panels), the number of estimators (y-axis) and the maximum tree depth (x-axis). We note that the highest achievable recall 75.5% is achieved with the smallest value LR=0.001, a maximum tree depth=4 and with n=100 individual estimators. Conceptually, this represents a comparatively small forest of comparatively shallow trees. Conveniently, such a model is also among the fastest for inference time given the relatively low complexity of the overall model.

**Figure S1.**
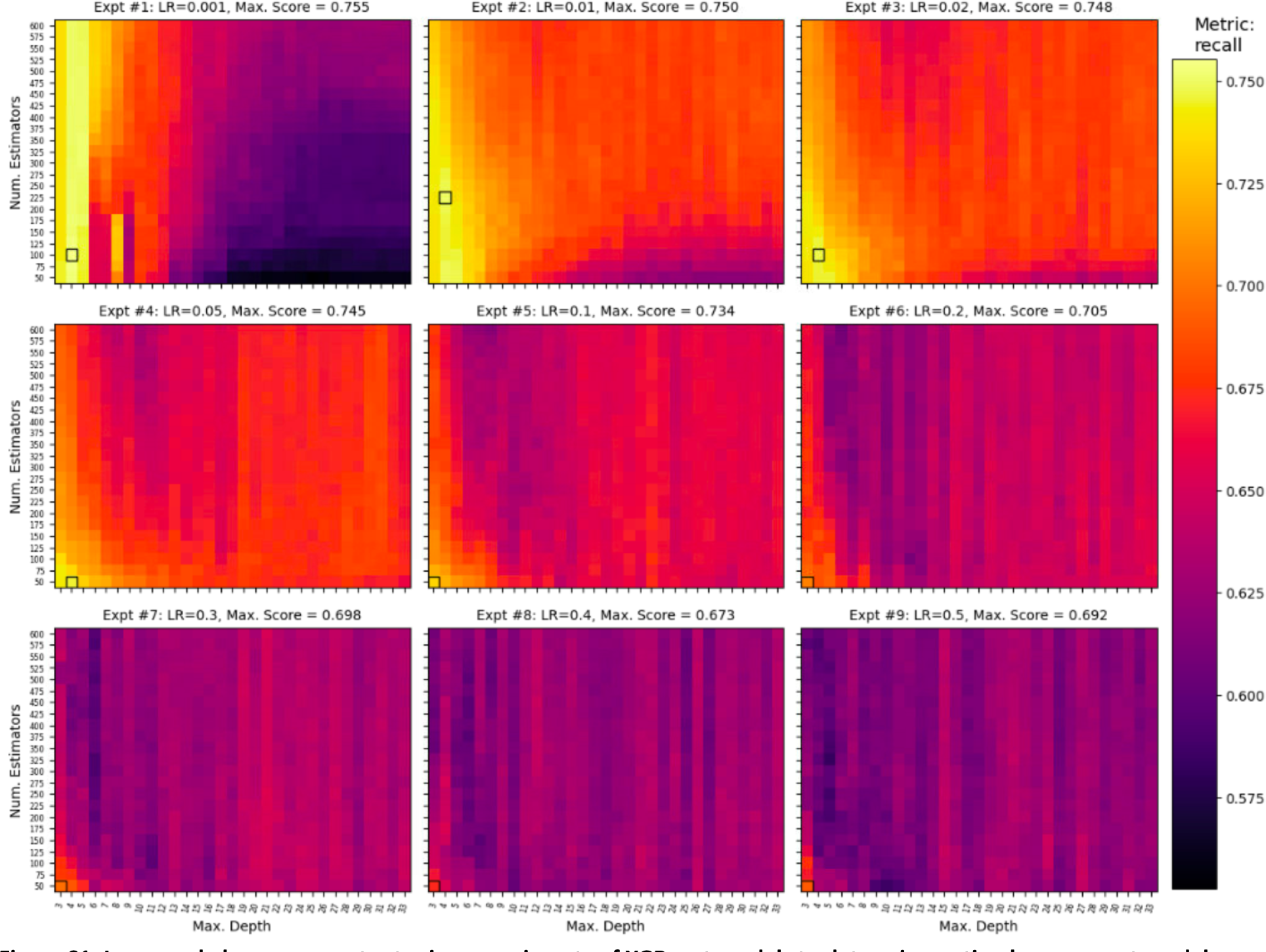
Large-scale hyperparameter tuning experiments of XGBoost models to determine op4mal component model configura4on. The individual heatmaps each represent one of nine learning rate values, and each varies the maximum tree (x-axis) and the number of component trees (y-axis) of each model. Top-performing models by Recall are identified within each learning rate (LR) panel for inter-panel comparison.

**Figure S2.**
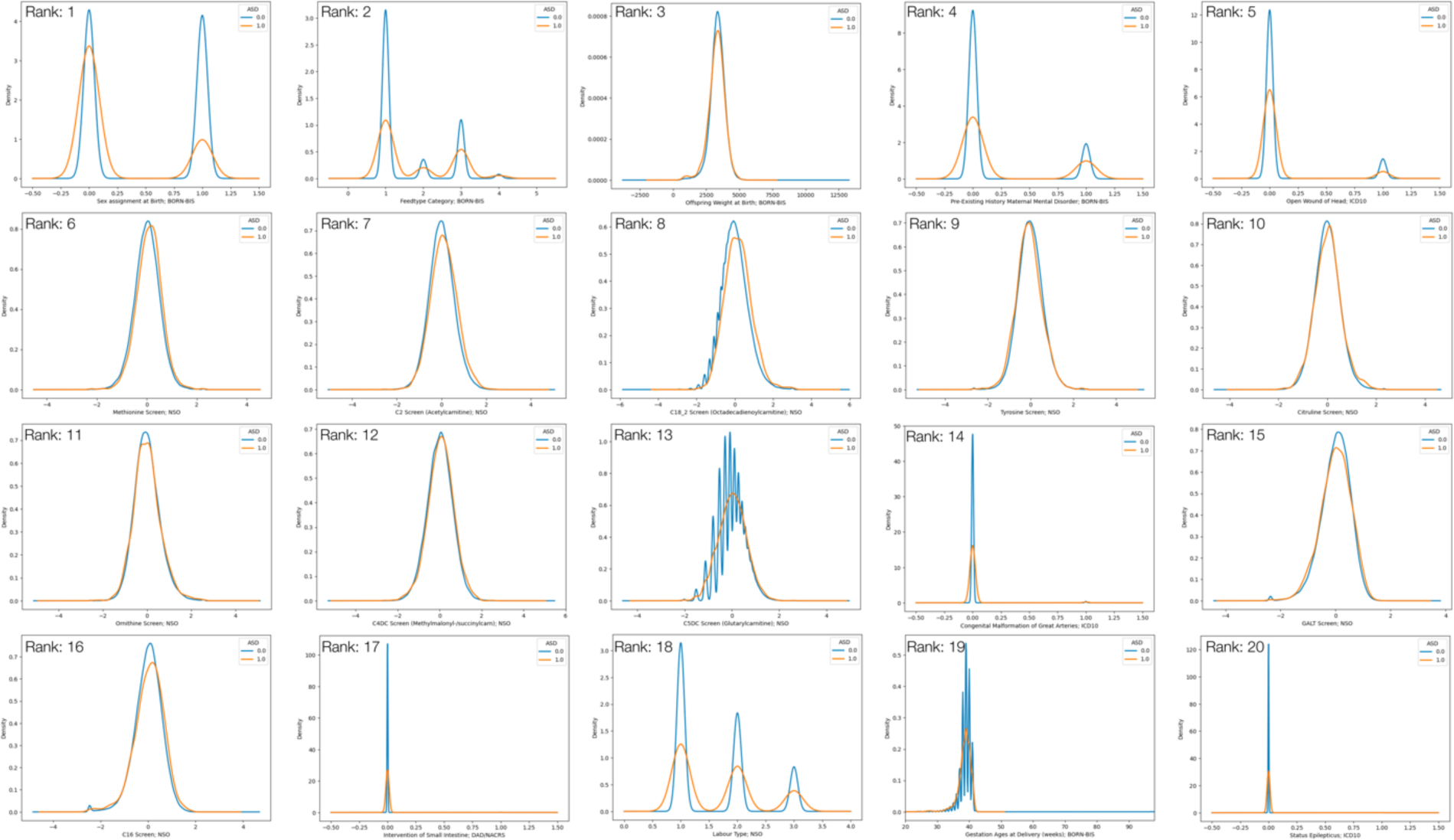
Kernel Density Es4ma4on Plots for the Top-20 Features from top-ranking XGBoost model SHAP Analysis.

**Figure S3.**
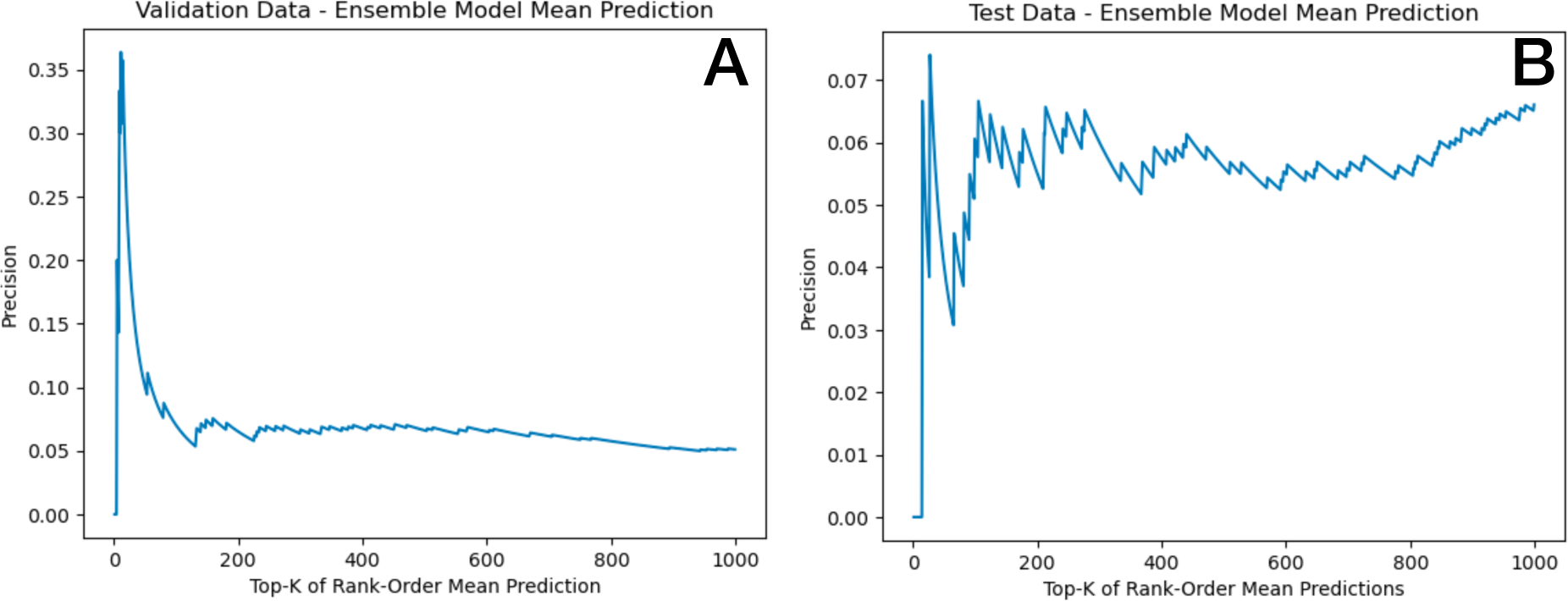
Top-K Precision by Rank Order of Mean Model Predic4on score.

**Table S3.**
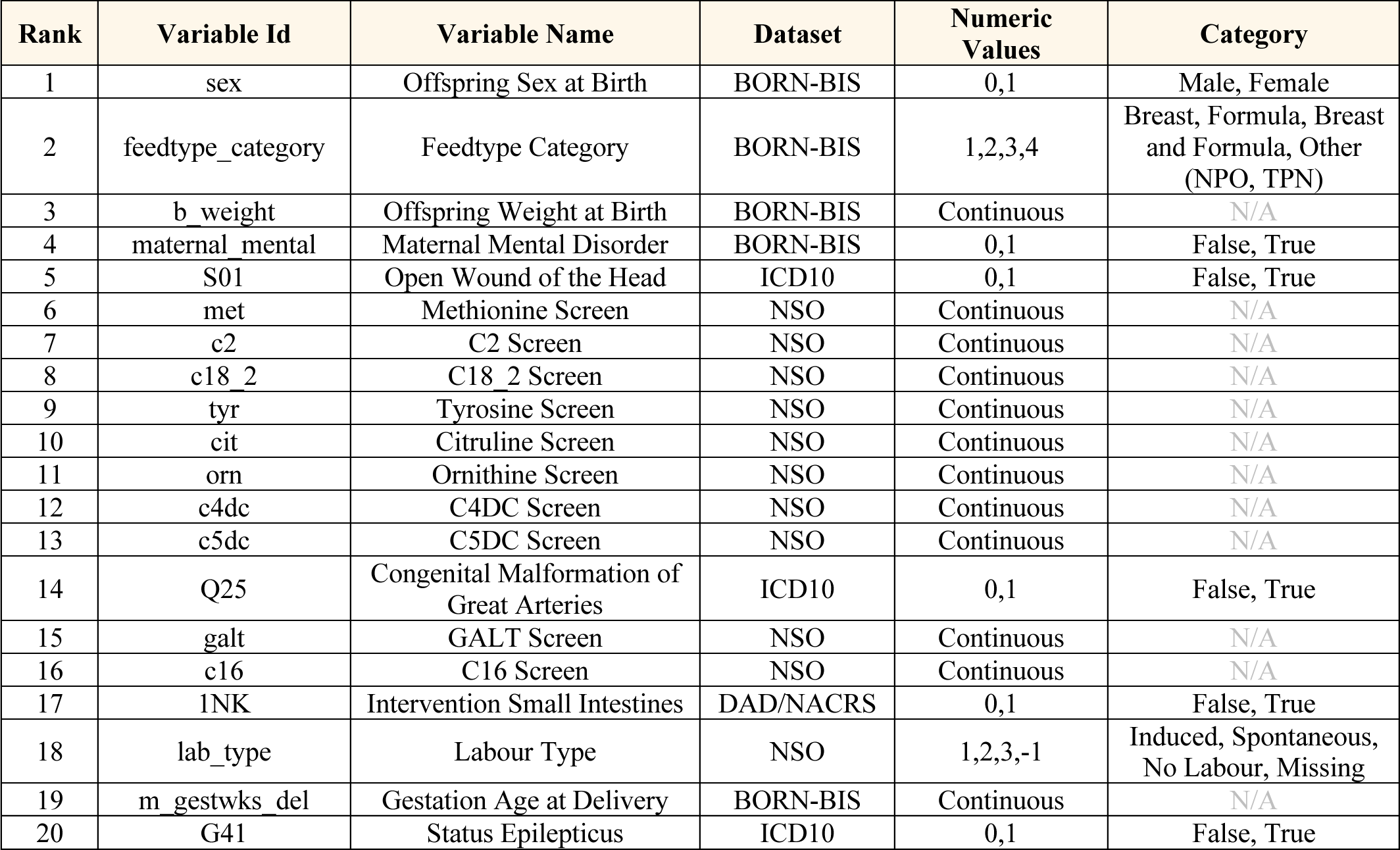
Tabula4on of the top-20 features.

